# Performance of empirical and model-based classifiers for detecting sucrase-isomaltase inhibition using the ^13^C-sucrose breath test

**DOI:** 10.1101/2024.05.01.24306704

**Authors:** Hannah Van Wyk, Gwenyth O. Lee, Robert J. Schillinger, Christine A. Edwards, Douglas J. Morrison, Andrew F. Brouwer

## Abstract

**Background:** Environmental enteric dysfunction (EED) is a syndrome characterized by epithelial damage including blunting of the small intestinal villi and altered digestive and absorptive capacity which may negatively impact linear growth in children. The ^13^C-sucrose breath test (^13^C-SBT) has been proposed to estimate sucrase-isomaltase (SIM) activity, which is thought to be reduced in EED. We previously showed how various summary measures of the ^13^C-SBT breath curve reflect SIM inhibition. However, it is uncertain how the performance of these classifiers is affected by test duration.

**Methods:** We leveraged SBT data from a cross-over study in 16 adults who received 0, 100, and 750 mg of Reducose, a natural SIM inhibitor. We evaluated the performance of a pharmacokinetic-model-based classifier, *ρ*, and three empirical classifiers (cumulative percent dose recovered at 90 minutes (cPDR90), time to 50% dose recovered, and time to peak dose recovery rate), as a function of test duration using receiver operating characteristic curves. We also assessed the sensitivity, specificity, and accuracy of consensus classifiers.

**Results:** Test durations of less than 2 hours generally failed to accurately predict later breath curve dynamics. The cPDR90 classifier had the highest area-under-the-curve and, by design, was robust to shorter test durations. For detecting mild SIM inhibition, *ρ* had a higher sensitivity.

**Conclusions:** We recommend SBT tests run for at least a 2-hour duration. Although cPDR90 was the classifier with highest accuracy and robustness to test duration in this application, concerns remain about its sensitivity to misspecification of CO_2_ production rate. More research is needed to assess these classifiers in target populations.

## Introduction

Environmental enteric dysfunction (EED) is characterized by atrophy of the small intestinal villi, resulting in increased intestinal permeability and nutrient malabsorption. It is thought to be highly prevalent among people in low- and middle-income countries who lack access to improved water, sanitation, and hygiene [1] and are therefore highly exposed to enteric pathogens [2,3]. EED is thought to play a central role in impaired linear growth (stunting) in infants and young children,[4] which impacts about 150 million children globally.

EED may be detected through the identification of histological features in small intestinal biopsies [5]. However, biopsies are invasive, require specialist skills and settings and are ethically questionable in sub-clinical EED, limiting the ability to accurately, efficiently, and inexpensively identify EED, especially in low-resource settings [6]. The most widely accepted non-invasive test, the lactulose:mannitol/rhamnose dual sugar urine absorption test, is time-consuming to administer and results may be inconsistent across laboratory platforms [7]. The ^13^C sucrose breath test (^13^C-SBT) has been proposed as an alternative [8]. The ^13^C-SBT is a stable-isotope breath test in which an individual ingests a dose of non-radioactive, ^13^C-labeled sucrose substrate, which is digested, absorbed, and metabolized, appearing on the breath as ^13^CO_2_. The ^13^C-SBT is intended to assess the activity of intestinal enzyme sucrase-isomaltase (SIM), a glucosidase enzyme that catalyzes the hydrolysis of carbohydrates [9]. Expression of SIM increases towards the tips of intestinal villi and therefore its activity is thought to be diminished in a damaged intestine [10-12]. Slower recovery of the tracer breath ^13^CO_2_ therefore indicates reduced gut enzyme metabolic function.

Although the ^13^C-SBT is attractive as a potential, non-invasive test of EED, it also has some limitations, which are common across ^13^C breath tests. Traditional measures used to interpret breath tests consist of empirically fitting a parametric curve to the percent dose recovery rate (PDRr) of ^13^CO_2_ on the breath, and calculating summary statistics, such as the cumulative percent dose recovered at 90 minutes (cPDR90), the time to peak PDRr (*T*_*peak*_), or the time to 50 percent dose recovered (*T*_50_) [13, 14]. However, these empirical measurements do not necessarily capture the underlying biological processes giving rise to the PDRr curve, and thus may be confounded by multiple aspects of the metabolism, some of which are unrelated to gut function. To address this concern we developed a mechanistic, pharmacokinetic model whose parameters represent the underlying biological processes occurring in the metabolism of the ^13^C-labeled sucrose tracer [15]. A model-based diagnostic *ρ* performed comparably to the highest-performing summary statistics in identifying experimentally induced sucrase-isomaltase inhibition in healthy adults [16].

In this analysis, we revisit these experiments to assess how the performance of the four highest performing classifiers, namely *ρ*, cPDR90, *T*_*peak*_ and *T*_50_, depend on the test duration. While experiments establishing and evaluating the SBT have used test durations of 4-8 hours [8, 15], there is a strong need to reduce the testing burden on participants, particularly for the target population of infants and children under 5 years. Additionally, because cPDR90, *T*_*peak*_, *T*_50_, and *ρ* capture different information about the breath curve, we will determine if consensus classifiers combining two or more classifiers can produce a more reliable diagnosis. In this research, we address these research gaps by assessing the accuracy of SBT curve projections based on shorter test duration, the performance of these three classifiers across test durations, and performance of consensus classifiers.

## Methods

### Data

The ^13^C-SBT breath curves used in this study were obtained in a crossover study conducted in Glasgow, United Kingdom, as previously described [16]. In brief, eighteen healthy adults were recruited to complete three breath test experiments under different experimental conditions designed to simulate different degrees of SIM inhibition. In this analysis, we only use data from the 16 participants who completed all three breath tests. The participants consisted of 8 female and 8 male participants with a mean age of 24.2 (SD= 5.0) and mean BMI of 24.5 (SD = 5.2). Participants were instructed to follow a low ^13^C diet for the three days preceding the experiments and to fast for eight hours prior to the test. In the first experiment, participants ingested 25 mg (0.84 mmol 13C) of highly enriched sucrose (≥99 atom% enriched; Sigma-Aldrich) to complete a baseline test. Breath samples were collected every 15 minutes for 4 hours into 12mL Exetainer breath-sample vials (Labco, United Kingdom). The relative difference in parts per thousand between the ratio *R*_*s*_ = [^13^C]/[^12^C] in the sample and the ratio (*R*_*std*_) of the laboratory CO_2_ standard (calibrated to the international calibration standard, VPDB, *R* =0.0112372) were determined by isotope ratio mass spectrometry (IRMS, AP-2003, Manchester, United Kingdom). Details on how this was converted to percent dose recovery rate are described in previous publications [15]. In the remaining experiments, participants were given in a random order either 100 and 750 mg of Reducose® (Phynova Group Ltd., Oxford, UK), a mulberry leaf extract (MLE) containing 5% 1-Deoxynojirimycin (an active α-glucosidase inhibitor) immediately prior to ingesting the 25 mg sucrose. Mulberry leaf extract has been shown to function as an intestinal SIM inhibitor, thus it is expected to induce similar ^13^CO_2_ excretion patterns to those that would be observed in patients with EED. The low dose of 100 mg Reducose was given to induce mild SIM inhibition, and the high dose of 750 mg was given to induce severe inhibition. Investigators received written informed consent from all participants and the study design was approved by the University of Glasgow College of Medical Veterinary and Life Sciences Research Ethics Committee (Application Number: 200190155).

### Mechanistic Model

In previous work [15], we developed a mechanistic, compartmental differential equation model that captured ^13^C-SBT breath curve dynamics and was practically identifiable, i.e., had parameters that could be uniquely estimated from data. In this model, the breath curve dynamics can be approximated as a combination of a gamma-distributed process with pharmacokinetic rate parameter *ρ*/2 and shape parameter 2 and an exponentially distributed process with rate parameter *πρ*. Because of the limitations of only observing the breath, the specific metabolic processes that these model processes represent are unknown *a priori*. In previous work [16], we demonstrated that both sucrase-isomaltase inhibition and the difference between fructose and glucose in the transport to and metabolism by the liver were reflected in the gamma-distributed process. In the model, we also account for the fraction of ^13^C that is exhaled, *κ*, as opposed to being secreted in urine or sequestered in adipose tissue.

When *π* ≠ 1, there is a closed-form solution for PDRr,

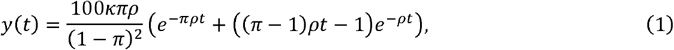

and the cPDR is given by

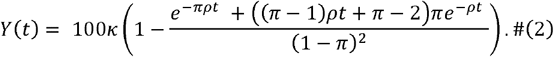

The classifiers we consider in this analysis are all obtained directly from the above equations: cPDR90 = Y(90), 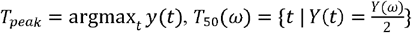, where *ω* is the test length, and *ρ* is the model-based classifier based on previous work [16]. Note that the definition of *T*_50_ used here, 50% of the cumulative percent dose recovered at test length *ω*, is different from previous work, [15] which defined it as time to recovery of 50% of the dose given. We use our definition here because most test participants do not recover 50% of the full dose over the testing period, especially in the case of mild-to-severe SIM inhibition.

### Parameter estimation

We estimated the parameter set *θ* = {*ρ, πρ,k*}corresponding to the best fit model by minimizing the negative log-likelihood (NLL), given by

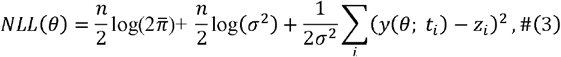

where *y* (*θ*; *t*_*i*_)is the value of the modeled PDR at time *t*_*i*_, *n* is the number of data points, 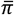 is the mathematical circle constant, *σ* is the standard deviation previously estimated to be 0.555 from best-fit curves [15], and *t*_*i*_ is the time at which measurement *z*_*i*_ was taken. In the case where the peak PDRr is not observed during the testing period, which was common among the 750 mg Reducose samples, *π ρ* and *k* are not identifiable. In this case, we added a penalty of size 0.1*k* onto the NLL to force the optimizer to select lower values of *k*. This forces the optimizer to choose larger values of *π ρ* that generate more realistic PDRr curves that do not extend over unrealistically long periods of time.

### Analytic Approach

The three objectives of this analysis were to 1) compare the accuracy of model projections as a function of test duration, 2) compare the performance of cPDR, *T*_*peak*_, *T*_50_, and *ρ*, as a function of test duration, and 3) assess the performance of consensus classifiers that combine two or more of the single classifiers. In this analysis, we examined test durations of 60, 90, 120, and 240 minutes. The following analysis plan outlines our approach:

1.) *Comparing model fits for 60-, 90-, 120-, and 240-minute duration tests*. For each participant j, we estimated 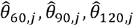, and 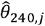, corresponding to the nine parameters that minimized the NLL for the baseline, 100 mg Reducose, and 750 mg Reducose breath curves, assuming that we only had the data from the first 60, 90, 120, and full 240 minutes, respectively. Then, to compare the model fits for the 60-, 90-, and 120-minute tests to the full dataset, we simulated the model for 240 minutes using each parameter set and calculated the NLL from each simulation against the full 240-minute data.
2.) *Comparing receiver operator characteristic (ROC) curves for ρ, cPDR, T*_50_ *and T*_*peak*_ *for 60-, 90-, 120-, and 240-minute duration tests*. We first noted that breath test curves that are initially slower (have a lower PDRr) also sustain a higher PDRr longer than the faster curves, allowing them to “catch up” to cumulative dose recovered of faster curves over time, which have a higher maximum PDRr, but a sharper curve around the peak. Therefore, the value of cPDR at a later time may be a less effective classifier than the value at an earlier time, and the cPDR with the highest dialogistic capability should be near the median *T*_*peak*_ . Thus, we first determined which cPDR classifier (cPDR60, cPDR90, cPDR120, or cPDR240) resulted in the most accurate classification using 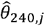 . As discussed in the results, we selected cPDR90. Then, we simulated the model for each parameter set *θ*_60,*j*_, *θ*_90,*j*_, *θ*_120,*j*_, and *θ*_240,*j*_, and estimated *ρ*, cPDR90, *T*_5O_ and *T*_*peak*_ in each case. We generated receiver operator characteristic (ROC) curves (which plot the true positive rate against the false positive rate as the classification threshold is varied) for all 12 combinations of test duration and classifier, for each of 4 groupings of the MLE experiments, corresponding to different clinical scenarios:
  1. Detection of *any* SIM inhibition (baseline versus *either* 100 or 750 mg MLE),
  2. Distinguishing between severe SIM inhibition vs none-to-mild (baseline or 100 mg MLE versus 750 mg MLE),
  3. Detection of mild SIM inhibition (i.e., baseline versus 100 mg MLE),
  4. Detection of severe SIM inhibition (baseline versus 750 mg MLE). The goal of the first two diagnostic groupings is to offer a single metric that captures the test’s ability to generate a binary diagnosis of SIM inhibition when the classifier takes any level of inhibition as an input, as would be the case in real-world applications. The last two classifiers assess the classifiers’ ability to identify differences in each of the three groups. For each ROC curve, we calculated the area under the curve (AUC) statistic, which represents the probability that a randomly selected positive sample is ranked as more likely to have SIM inhibition than a randomly selected negative sample [17].
3.) *Assessment of single and consensus classifiers*. We assessed the accuracy, sensitivity, specificity, and Matthew’s correlation coefficient (MCC) of each classifier at their optimal thresholds (the cutoff threshold that maximizes the sum of the sensitivity and specificity of the test [18]). The MCC is an alternative accuracy measurement that is preferred for unbalanced datasets and has a range of [−1,1] where 1 means perfect classification, 0 corresponds to a coin toss classifier, and −1 is perfect misclassification [19]. We further examined the accuracy, sensitivity, specificity, and Matthew’s correlation coefficient (MCC) of consensus classifiers consisting of each combination of the individual metrics *ρ*, cPDR90, *T*_50_, and *T*_*peak*_ at their optimal thresholds. To generate these statistics for each participant in each experiment, we generated consensus diagnoses for each participant based on each combination of the individual classifiers. For example, assuming that a positive diagnosis of SIM inhibition is defined by *both ρ* and cPDR90 (*ρ* ∩ cPDR90) indicating inhibition or assuming that a positive diagnosis is defined by *either ρ* and cPDR90 (*ρ* ∪ cPDR90) indicating inhibition. We assessed this for each possible combination of three classifiers at a time. For example, for *ρ*, cPDR90, and *T*_50_ that is: *ρ* only, cPDR90 only, *T*_50_ only, *ρ* ∩ cPDR90, *ρ* ∩ *T*_50_, cPDR90 ∩ *T*_50_, *ρ* ∩ cPDR90 ∩ *T*_50_, *ρ* ∪ cPDR90, *ρ* ∪ *T*_50_, cPDR90 ∪ *T*_50_, and a majority rules classifier. For the majority rules classifier, a positive diagnosis was generated if at least two of the individual classifiers are positive. To compare consensus classifier performances for each of the three MLE doses, we generated this result for each of the same four comparison groups outlined in step 2. We repeated this for the 60-, 90-, and 120-minute test lengths to assess classifier robustness to decreased data.

## Results

### Comparing model fits for 60-, 90-, 120-, and 240-minute tests

Projections from fitting the model only to the first 60 minutes of the data were consistently poor fits for the later data (illustrative examples given in Fig. 1a, with full results in Fig S1 in the SI appendix). For the 60-min test duration, random variations present in each data point had a higher influence on the model fit than it did with longer test periods, causing model trajectories in hours 1–4 to be heavily impacted by these fluctuations. Additionally, the inability to observe the peak PDRr in the first hour—particularly for the 750 mg group—meant that *πρ* and *κ* were unidentifiable at this test duration, severelylimiting the model’s inferential ability for later hours. While the 90-minute test duration generally improved the fit somewhat, the improvement was not consistent across participants, and many curves fit to 90 minutes were poorly predictive of later dynamics. When comparing the NLLs between the models fit to data from each test length (Figure 1b), we found substantial heterogeneity in the impact of test length on model fit, depending on the participant. The fits at shorter tests lengths were typically better in participants for whom the peak PDRr was reached within the respective test length (see Fig S1 in the SI appendix). In general, the projections from curve fit to the data from the first 120 minutes are very similar to the curves fit to the full data, with some outliers. In the following sections, we assessed how the improvement in model fit is reflected in the diagnostic capability of the test.

**Figure 1:**
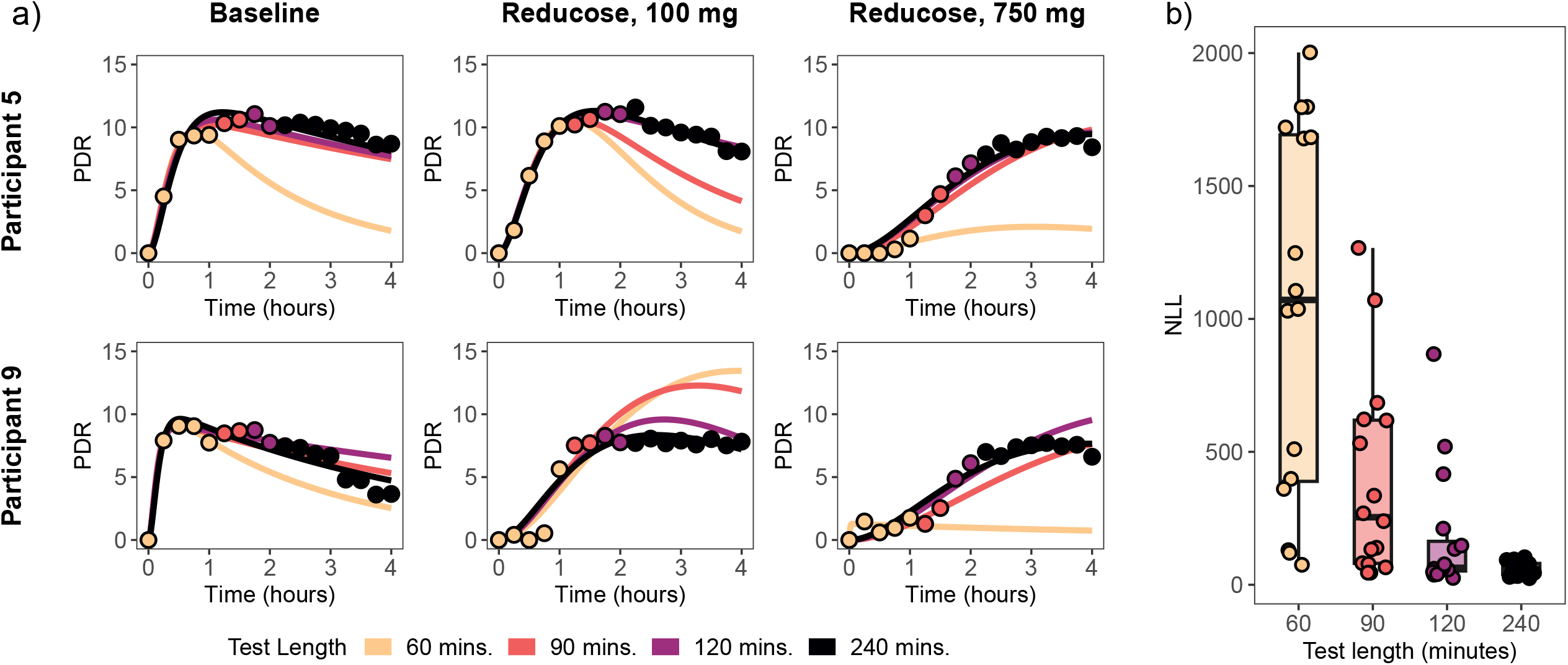
a) Model best fits for 60-, 90-, 120-, and 240-minute test durations for two study participants b) Boxplot of negative log-likelihoods (NLLs), a measure of how well the model fits the data, for each test duration, with larger values indicating poorer fit. Plots for all participants are given in Fig S1 in the SI appendix.

We also plot the value of each classifier for each participant and test duration across the three MLE doses to visualize each classifier’s sensitivity to MLE dosage (Fig. 2). The plots for cPDR90 (Fig. 2a) show that this classifier has the strongest distinction between the lowest two doses (i.e., baseline or 100 mg MLE) and the 750 mg dose; however, the distinction between the baseline and 100 mg MLE dose is minor. By contrast, the figure for *ρ* (Fig. 2b) shows a better separation between the value of *ρ* and MLE dose, indicating that this classifier may be more sensitive to detecting lower MLE doses, which represent mild SIM inhibition.

**Figure 2:**
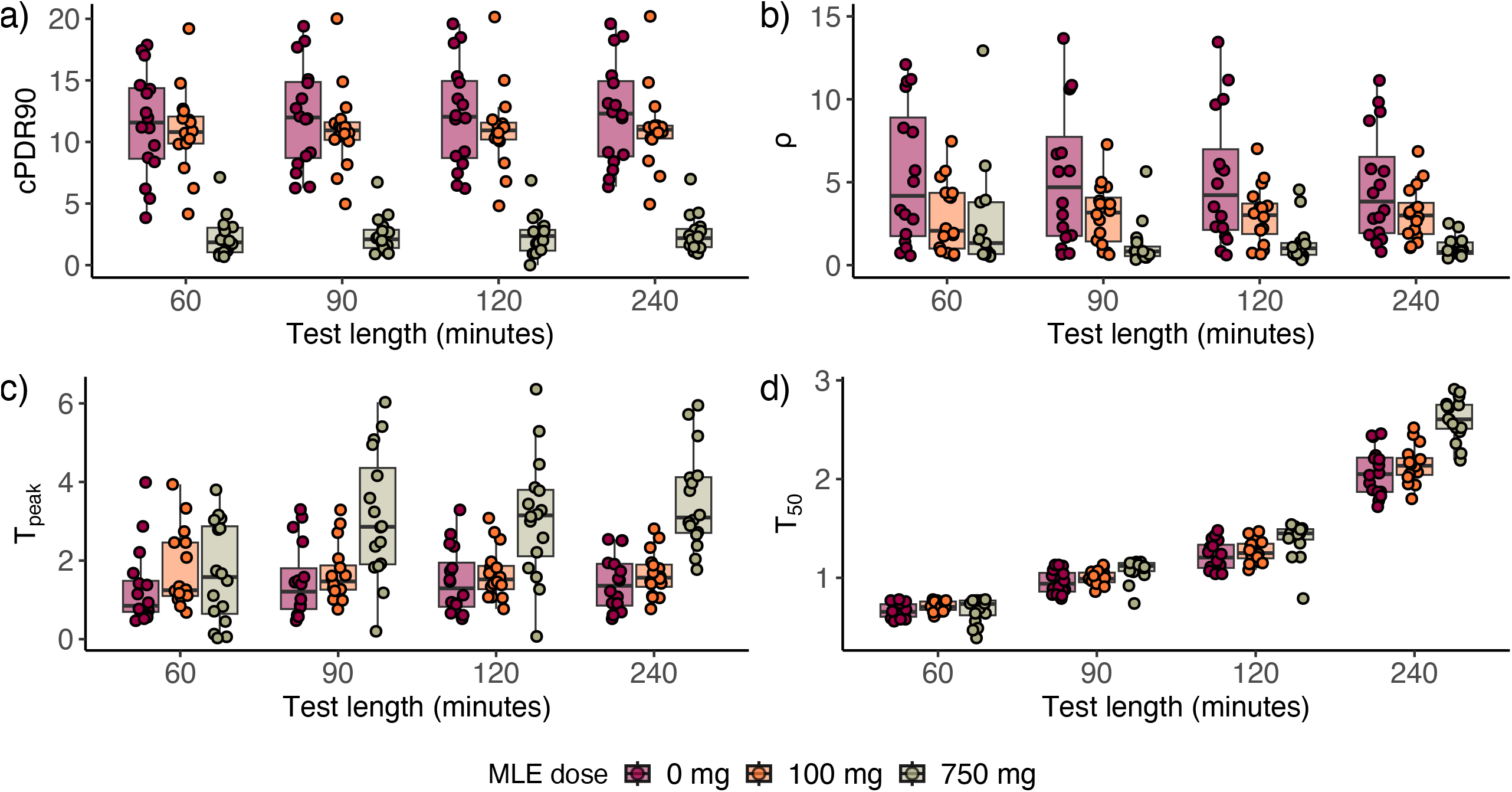
Classifier values for 60-, 90-, 120-, and 240-minute ^13^C-sucrose breath test durations for baseline, 100, and 750 mg doses of Reducose®, a mulberry leaf extract (MLE) that acts as a sucrase-isomaltase inhibitor for (a) cPDR90, (b) *ρ*, (c) *T*_*peak*_, and (d) *T*_50_.

### *Comparing ROC curves for ρ, cPDR, time to 50% dose recovered (T*_50_*), and time to peak (T*_*peak*_*) for 60-, 90-, 120-, and 240-minute duration tests*

We found that cPDR90 and cPDR60 outperformed cPDR120, and cPDR240 in ROC curves (Fig. S3). Prior literature has used cPDR90, so, for consistency, we selected cPDR90 as the cPDR classifier to compare to *ρ, T*_50_ and *T*_*peak*_. Our ROC curves for baseline versus either 100 or 750 mg MLE (Fig 3, blue) and baseline or 100 mg MLE versus 750 mg MLE (Fig 3, yellow) showed that cPDR90 had the highest AUC for each test length and comparison group. The cPDR90 classifier also maintained the same AUC (0.99) for each test length for 0 or 100 mg v. 750 mg and only saw a slight decrease in the AUC for the other comparison group (0.79 at 240 minutes versus 0.77 at 60 minutes). The ROC curves corresponding to baseline versus 100 mg (Fig S2) show that *ρ* outperforms cPDR90 for distinguishing mild SIM inhibition from none (AUC ranges: 0.61-0.66 for *ρ* and 0.55-0.60 for cPDR90). However, because *ρ* was not as accurate at distinguishing severe inhibition from no inhibition in these data (AUC range: 0.58-0.93), its AUC is always below the AUCs corresponding to cPDR90 in Fig 3. Additional ROC curves assuming the data is available at 15 min for hours 0-1, every 30 min for hours 1-4 is available in the SI appendix as an additional sensitivity analysis (Fig. S4).

**Figure 3:**
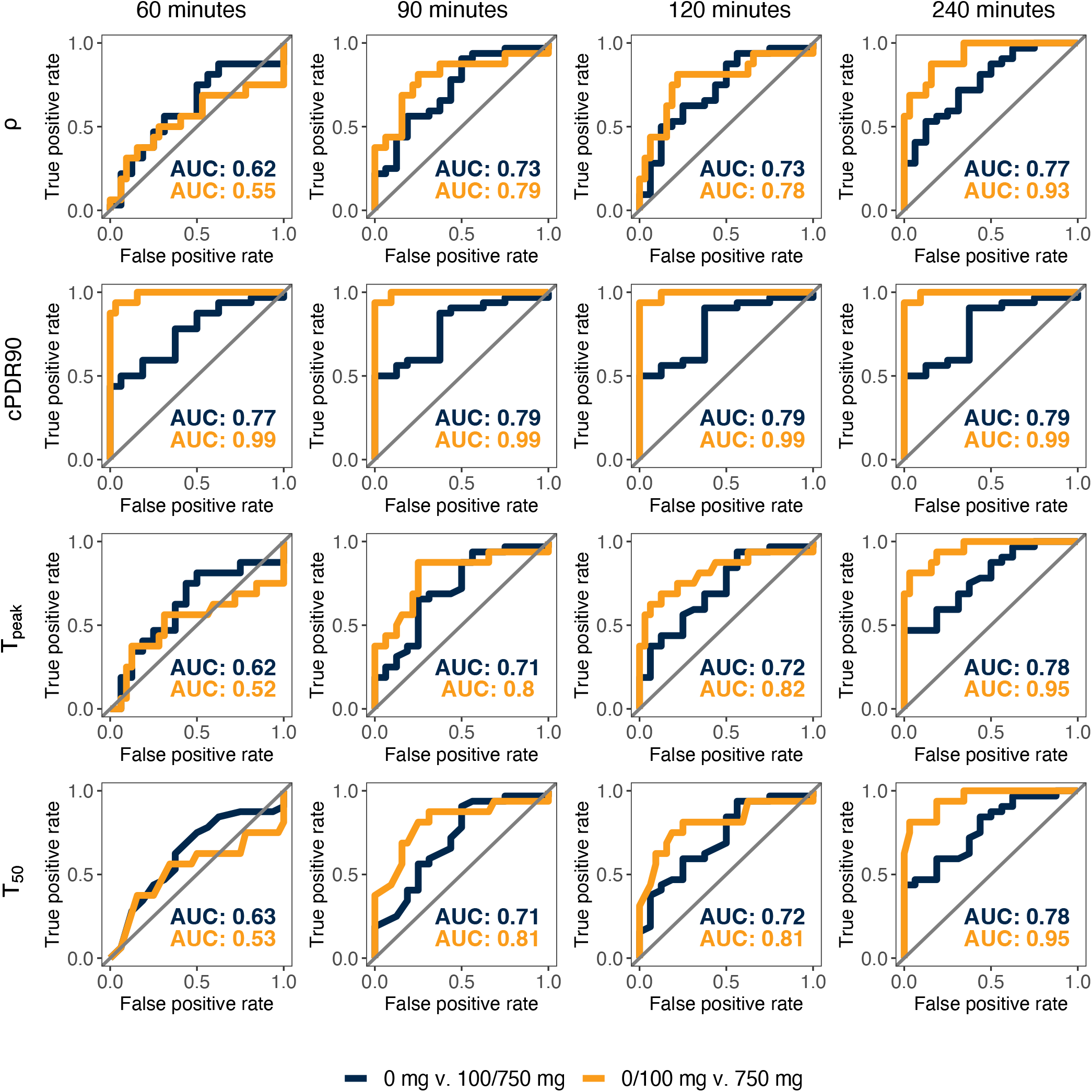
ROC curves for 60-, 90-, 120-, and 240-minute ^13^C-sucrose breath test durations for baseline versus either 100 or 750 mg doses of Reducose®, a mulberry leaf extract (MLE) that acts as a sucrase-isomaltase inhibitor (blue), and baseline or 100 mg MLE versus 750 mg MLE (orange).

### Assessment of consensus classifiers

Table 1 shows the results of the consensus classifiers including cPDR90, *ρ*, and *T*_50_, which were the three highest performing classifiers according to Fig. 2. The consensus classifiers including *T*_*peak*_ are available in the Supplementary Material, (Tables S1 through S4). Consistent with the results from the ROC curves, the performance statistics of the consensus classifiers (Table 1) show that cPDR90 alone has the highest accuracy and MCC for each of the four Reducose dose comparison groupings. However, for sensitivity, cPDR90 is outperformed by *ρ* and *T*_50_ for the baseline versus 100 mg group, and by *ρ* ∪ cPDR90 for 0 versus 750 mg and 0/100 versus 750 mg. For the shorter test durations, cPDR90 continues to be the best classifier for all comparison groups for the 120-minute test length (Table S1). However, *ρ* and *T*_50_ surpass cPDR90 by the 90- and 60-minutes lengths for the baseline versus 100 mg and 0 v 100/750 mg comparison groups (Tables S1 and S2). The consensus classifiers also perform better than the individual classifiers at these shorter test durations. For example, at the 60-minute test duration, cPDR ∩ *T*_*peak*_ and *ρ* ∩ cPDR ∩ *T*_*peak*_ had the highest accuracy and MCC for the baseline versus 100 mg group (Table S1).

**Table 1:** Accuracy, sensitivity, specificity, and Matthew’s Correlation Coefficient (MCC) of consensus metrics for the 240-minute duration test.

| | $\rho$ | cPDR | $T_{50}$ | $\rho \cap$<br>cPDR | $\rho \cap$<br>$T_{50}$ | cPDR<br>$\cap T_{50}$ | $\rho \cup$<br>cPDR ∪<br>$T_{50}$ | $\rho \cup$<br>cPDR | $\rho \cup$<br>$T_{50}$ | cPDR<br>$\cup T_{50}$ | Majority<br>rules |
| --- | --- | --- | --- | --- | --- | --- | --- | --- | --- | --- | --- |
| **Accuracy** |  |  |  |  |  |  |  |  |  |  |  |
| 0 v. 100 mg | 0.66 | **0.72** | 0.66 | **0.72** | 0.66 | **0.72** | 0.66 | 0.66 | 0.66 | 0.66 | 0.66 |
| 0 v. 750 mg | 0.88 | **0.97** | 0.91 | 0.91 | 0.91 | 0.91 | 0.94 | 0.94 | 0.88 | **0.97** | 0.91 |
| 0/100 v. 750 mg | 0.85 | **0.98** | 0.92 | 0.94 | 0.92 | 0.94 | 0.90 | 0.9 | 0.85 | 0.96 | 0.92 |
| 0 v. 100/750 mg | 0.71 | **0.81** | 0.62 | 0.73 | 0.62 | 0.62 | 0.79 | 0.79 | 0.71 | **0.81** | 0.73 |
| **Sensitivity** |  |  |  |  |  |  |  |  |  |  |  |
| 0 v. 100 mg | **0.94** | 0.81 | **0.94** | 0.81 | **0.94** | 0.81 | **0.94** | **0.94** | **0.94** | **0.94** | **0.94** |
| 0 v. 750 mg | 0.88 | 0.94 | 0.81 | 0.81 | 0.81 | 0.81 | **1.00** | **1.00** | 0.88 | 0.94 | 0.81 |
| 0/100 v. 750 mg | 0.88 | 0.94 | 0.81 | 0.81 | 0.81 | 0.81 | **1.00** | **1.00** | 0.88 | 0.94 | 0.81 |
| 0 v. 100/750 mg | 0.72 | **0.91** | 0.44 | 0.72 | 0.44 | 0.44 | **0.91** | **0.91** | 0.72 | **0.91** | 0.72 |
| **Specificity** |  |  |  |  |  |  |  |  |  |  |  |
| 0 v. 100 mg | 0.38 | **0.63** | 0.38 | **0.63** | 0.38 | 0.62 | 0.38 | 0.38 | 0.38 | 0.38 | 0.38 |
| 0 v. 750 mg | 0.88 | **1.00** | **1.00** | **1.00** | **1.00** | **1.00** | 0.88 | 0.88 | 0.88 | **1.00** | **1.00** |
| 0/100 v. 750 mg | 0.84 | **1.00** | 0.97 | **1.00** | 0.97 | **1.00** | 0.84 | 0.84 | 0.84 | 0.97 | 0.97 |
| 0 v. 100/750 mg | 0.69 | 0.63 | **1.00** | 0.75 | **1.00** | **1.00** | 0.56 | 0.56 | 0.69 | 0.62 | 0.75 |
| **MCCs** |  |  |  |  |  |  |  |  |  |  |  |
| 0 v. 100 mg | 0.38 | **0.45** | 0.38 | **0.45** | 0.38 | **0.45** | 0.38 | 0.38 | 0.38 | 0.38 | 0.38 |
| 0 v. 750 mg | 0.75 | **0.94** | 0.83 | 0.83 | 0.83 | 0.83 | 0.88 | 0.88 | 0.75 | **0.94** | 0.83 |
| 0/100 v. 750 mg | 0.69 | **0.95** | 0.81 | 0.86 | 0.81 | 0.86 | 0.80 | 0.80 | 0.69 | 0.91 | 0.81 |
| 0 v. 100/750 mg | 0.39 | **0.56** | 0.45 | 0.45 | 0.45 | 0.45 | 0.51 | 0.51 | 0.39 | **0.56** | 0.45 |

## Discussion

In this analysis, we leveraged a mechanistic model to compare the performance of traditional, empirical classifiers (i.e., cPDR90, *T*_50_ and *T*_*peak*_) of ^13^C-SBT breath test to that of a mechanistic, pharmacokinetic model-based classifier. We found that, under typical data variation, 60-minute duration tests were insufficient to adequately project breath trajectories, primarily due to limited ability to observe some of the post-peak PDRr trajectory in these time lengths (Figure 1). Thus, we recommend ^13^C-SBT future protocols use a 120-minute or longer test duration. For the ^13^C-SBT, test durations up to 240 minutes saw enhanced accuracy and improvement in the performance of the *T*_50_, *T*_*peak*_ and model-based classifier, but the ability to estimate SIM activity from a shorter-duration test supports the wider use of the ^13^C-SBT for gut dysfunction research and, potentially, for future clinical usage. However, other ^13^C breath tests may have different recommended durations if the distribution of peak PDRr is different for a different isotopic tracer, so further study of potential tracers could identify a substrate with a further reduced testing burden.

Our results from the classifier performance comparison show that cPDR90 was the best classifier (by AUC) at each test length, compared to *ρ, T*_50_, and *T*_*peak*_ (Fig. 2). These results suggest that, even though cPDR is not directly measuring the underlying biological mechanisms, slow cumulative recovery of the breath is highly informative. We also found that the consensus classifiers generally performed worse than the individual ones, largely because cPDR90 was highly accurate on its own for this population. However, as we see in Eq 2, the cumulative percent dose recovery is highly dependent on κ, the fraction of tracer that is excreted through the breath. Hence, the performance of cPDR will be highly sensitive to variations in this fraction or, as we previously showed [15], to misestimation of the production rate of C0_2_, V_CO2_, which is estimated based on body size. As a result, associations between cPDR and demographic or anthropometric variables may be introduced through differential bias In V_CO2_ estimates. This limits the application of the cPDR to the ^13^C-as a test of EED in young children, because poorer growth is posited to be a key consequence of EED. We will explore anthropometric and demographic associations with breath curve dynamics in future work). Hence, we caution against taking our results as evidence that cPDR90 is the only classifier needed. Additionally, we note that both *ρ* and *T*_*peak*_ outperform cPDR90 for model sensitivity (Table 1) and for distinguishing the 100 mg dose from baseline (Fig. S2). Currently, it is unknown whether SIM inhibition in typical a case of EED is more similar to the inhibition induced by the 100 mg MLE dose or the 750 mg dose.

We found that some classifiers were quite accurate at shorter test lengths or even had a higher AUC at shorter test lengths. For example, the *T*_*peak*_ AUC for 0 mg v. 100 mg has a higher AUC (0.68) when generated from the 60-minute data as opposed to the 240-minute data (AUC = 0.61). However, this result does not necessarily indicate that those classifiers were robust to a shorter test length. Rather, this behavior is a data artifact: the curves estimated at the shorter test lengths are often poor fits to the full breath curve (Fig S1), and thus they happen to have better classifier performance only by accident. The same classifier might perform drastically worse on a different dataset for that test duration. This phenomenon is not a limitation of our analysis but a limitation of short-duration breath tests, and it has implications for future studies. Participants do not always complete the full breath collection protocol, but researchers may want to include the data that were collected. We advise having a clear exclusion criterion in ^13^C-SBT studies for participants who do not complete at least 90-min of breath collection.

The primary strength of this study is the crossover study design. The experimental design artificially induced SIM inhibition in the study participants, making the comparison between experiments unconfounded by other factors that would be likely present in cases and controls from separate populations. However, because the data is from healthy adult participants for whom SIM was experimentally inhibited, the performance of the classifiers may be different from the target population, i.e., children in low-resource settings, which means that the external generalizability may be limited. In addition, the small samples size makes the results more sensitive to random measurement error. For the ^13^C-SBT to move from being a specialized research tool to wider useability, further research that includes a larger sample size and inclusion of study participants from the target population will be needed. Our results facilitate this work by suggesting a shortened, 120-minute test duration, that may be more feasible for infants and young children compared to the prior, standard 4-hour test.

## Conclusion

We assessed the performance of two empirical classifiers, cPDR90, *T*_50_, and *T*_*peak*_, and one model-based classifier, *ρ* for the SBT over different test lengths. Based on curves fit to different test lengths, we recommend that ^13^C-SBT protocols include 120-min or longer test durations and that participants who collect less than 90 min of breath be excluded. We found that, overall, cPDR90 was the most accurate classifier in these data; however, limitations of this classifier include uncertainty around its performance in the target population and lower sensitivity in detecting cases of mild SIM inhibition. The model-based classifier *ρ* addresses both concerns because it is more reflective of the underlying biological processes giving rise to the PDRr curves. We recommend multiple classifiers continue to be considered in future work assessing the performance of the ^13^C-SBT as a diagnostic test of EED or other dysfunctions that reduce SIM activity.

## Data Availability

Data described in the manuscript (Data_MLE.csv) are made publicly and freely available without restriction at https://doi.org/10.5281/zenodo.8387995.

## Author contributions

Conceptualization (of this analysis): AFB, GOL, DJM; Methodology: AFB, HVW; Investigation: RJS, CAE, DJM; Formal Analysis: HVW; Visualization: HVW; Writing - Original Draft: HVW; Writing - Review & Editing: AFB, GOL, HVW, DJM. Supervision: DJM (lab), AFB (analysis). All authors read and approved the final manuscript.

## Acknowledgements

This project was funded through the International Atomic Energy Agency (IAEA) coordinated research projects E4.10.16 and E430336, United States National Science Foundation (NSF) grant DMS1853032, and United States National Institutes of Health (NIH) grant K01AI145080. The NSF and NIH were not involved in study design; collection, analysis, and interpretation of data; writing of the report. The IAEA was involved in study design of the data collection. We also thank Dr. Mamane Zeilani, Nutriset for part-funding of this work and Dr. Andrew Gallagher of Phynova Group Ltd for the supply of Reducose for this study. The industry collaborators had no role in study design, collection, analysis, interpretation of the data or writing of the report.

## Data Availability Statement

