## Supplementary Material for "Performance of empirical and model-based classifiers for detecting sucrase-isomaltase inhibition using the ^13^C-sucrose breath test"

### Supplemental materials

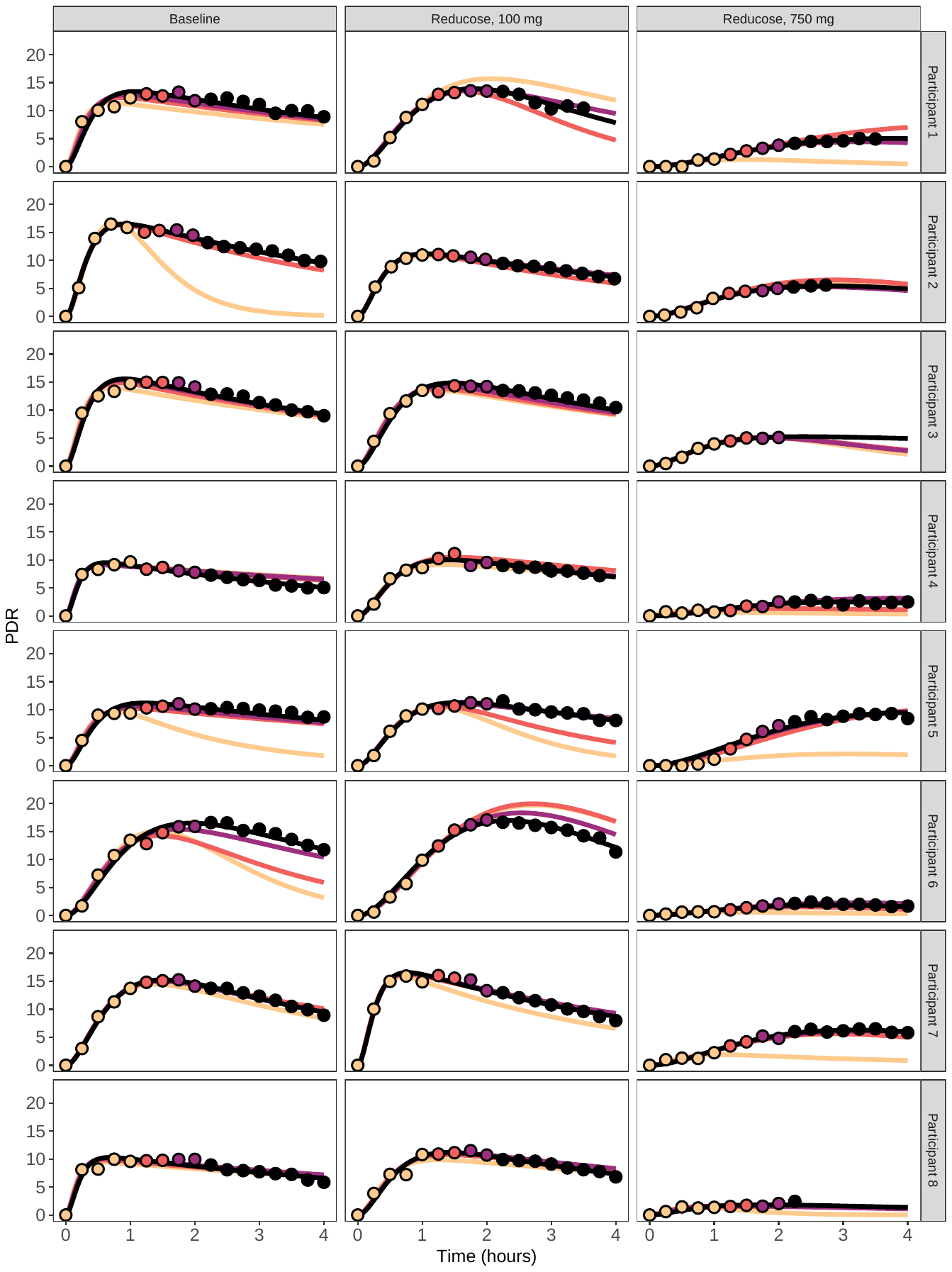

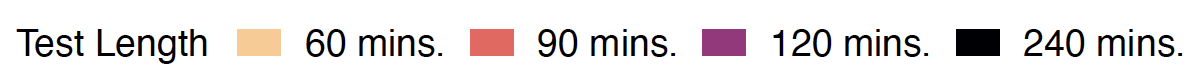

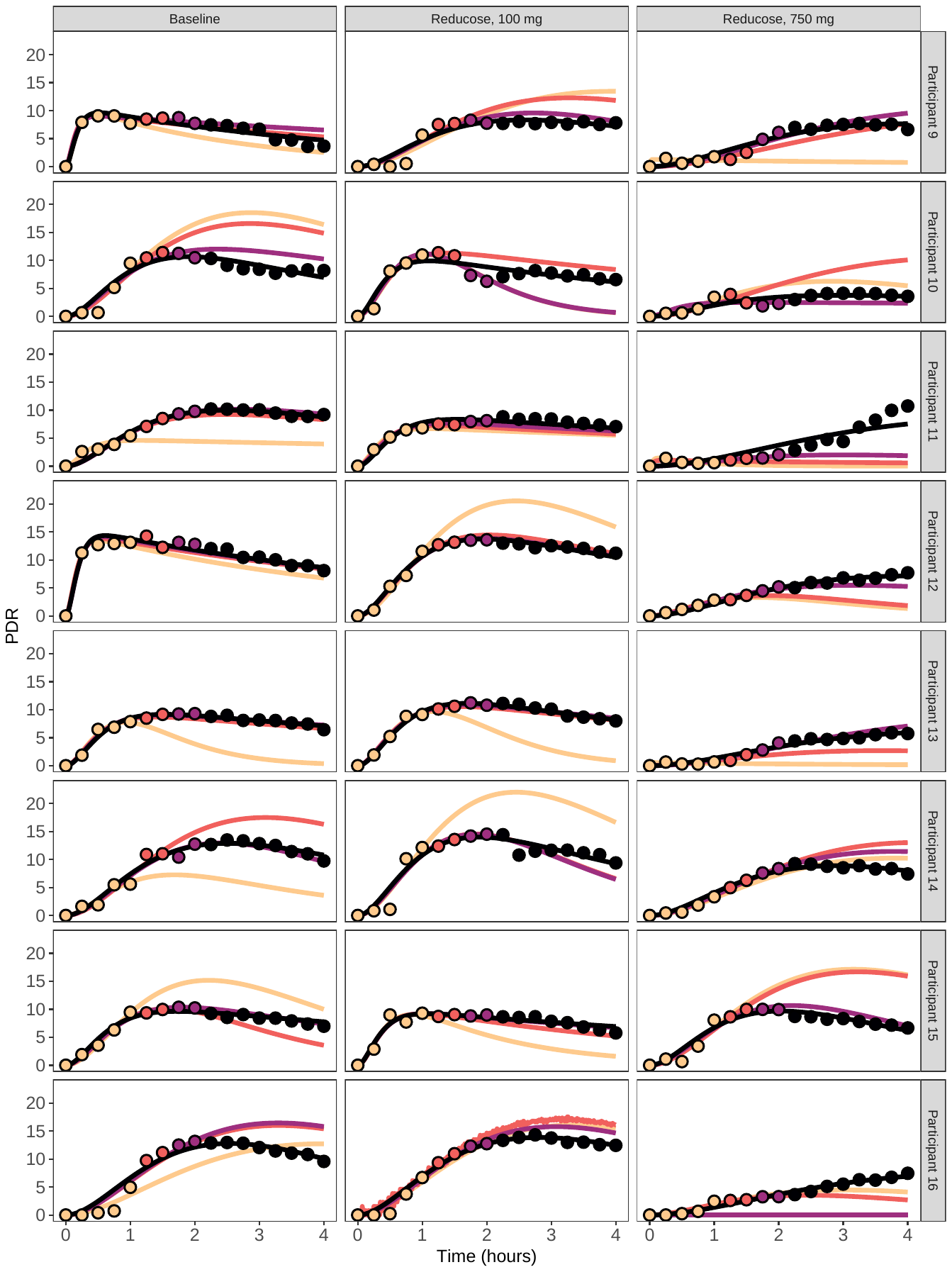

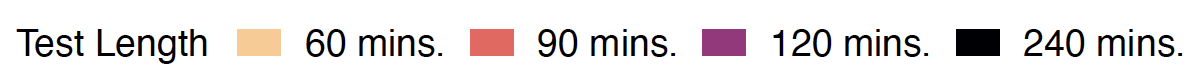

**Figure S1**: Data and model projections when fit to the first 60, 90, 120, or 240 minutes of test data.

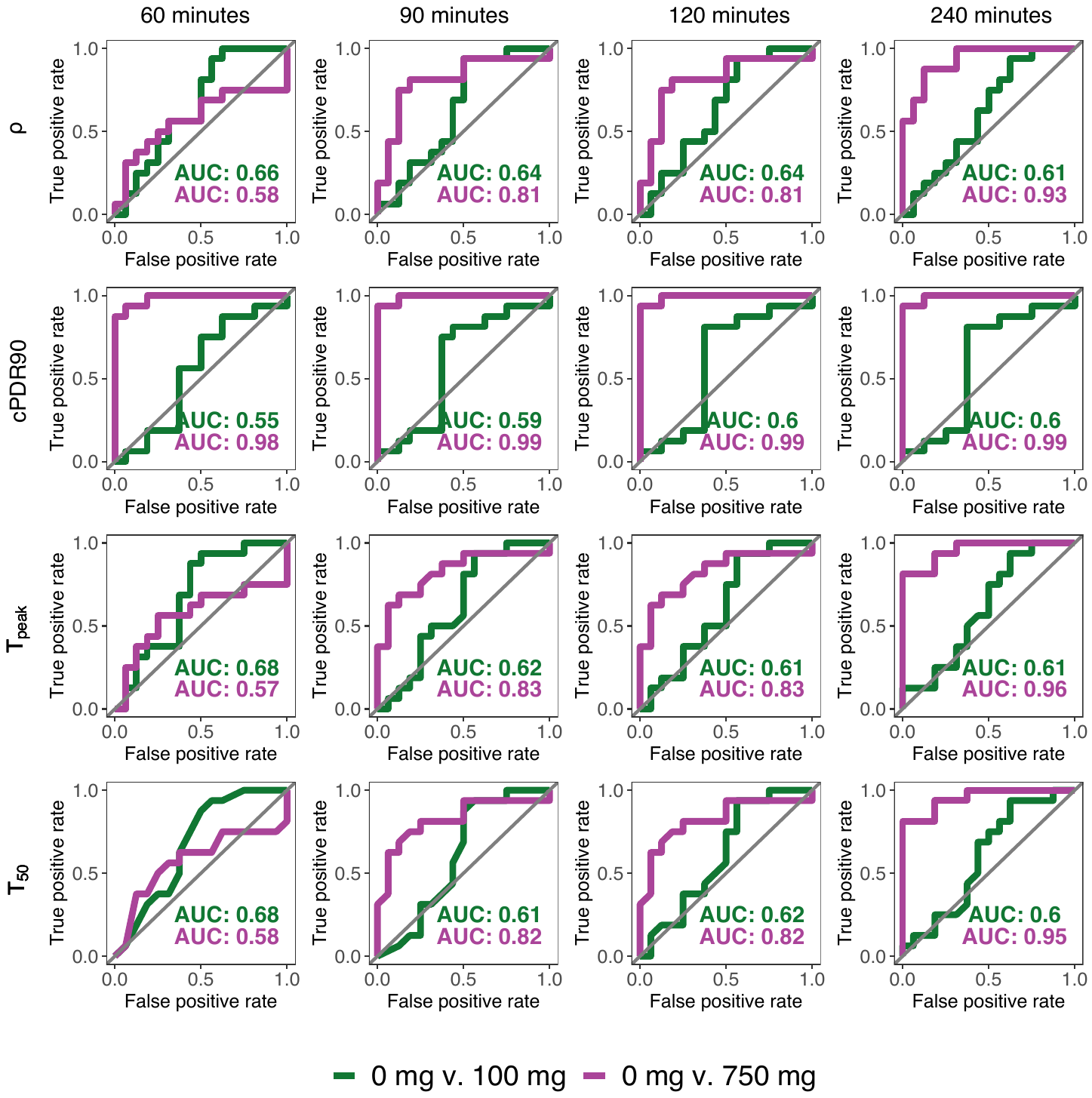

**Figure S2:** ROC curves for data up to 60-, 90-, 120-, and 240-minute test durations.

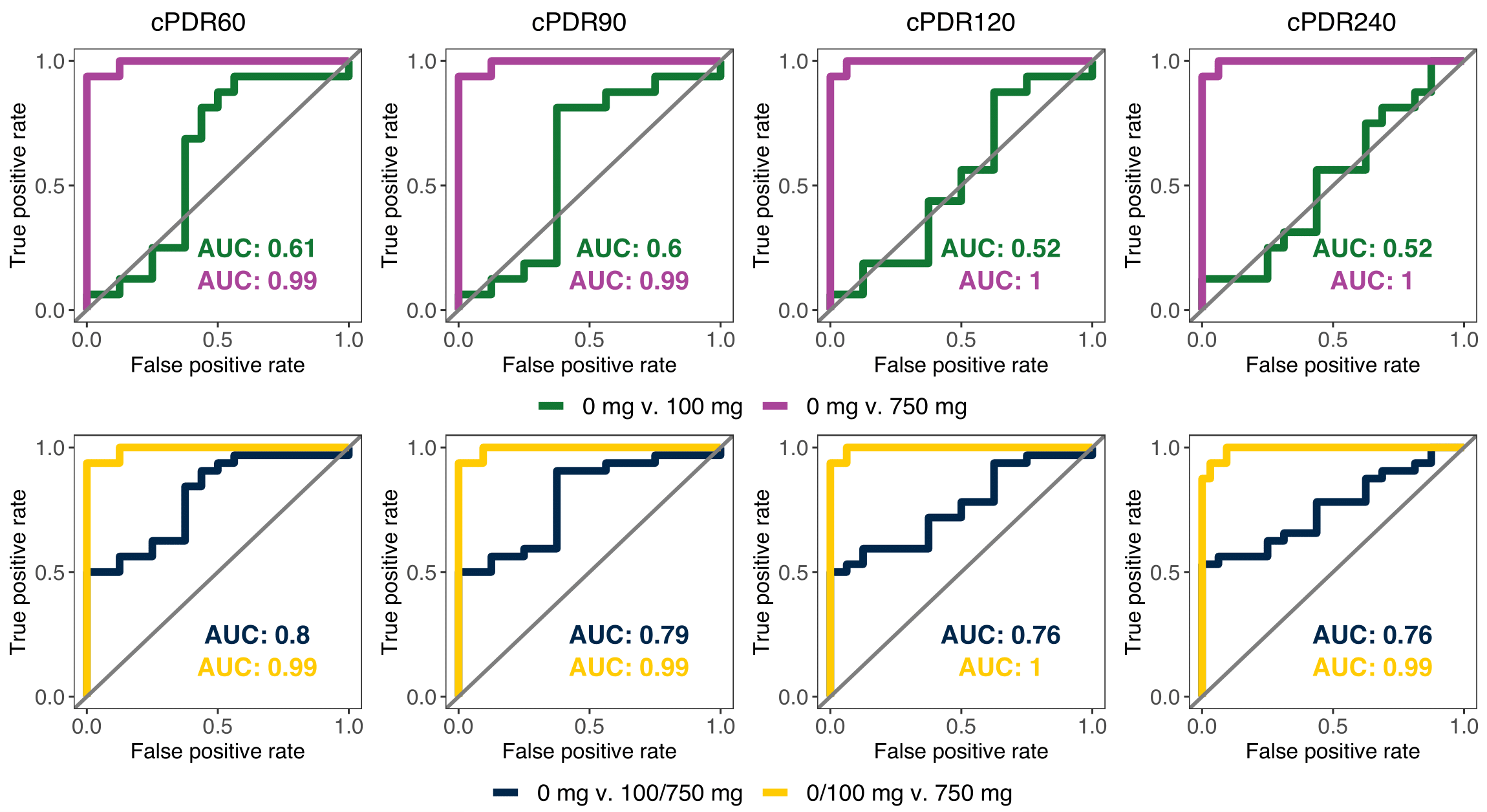

**Figure S3:** ROC curves for cPDR60, cPDR90, cPDR120, and cPDR240.

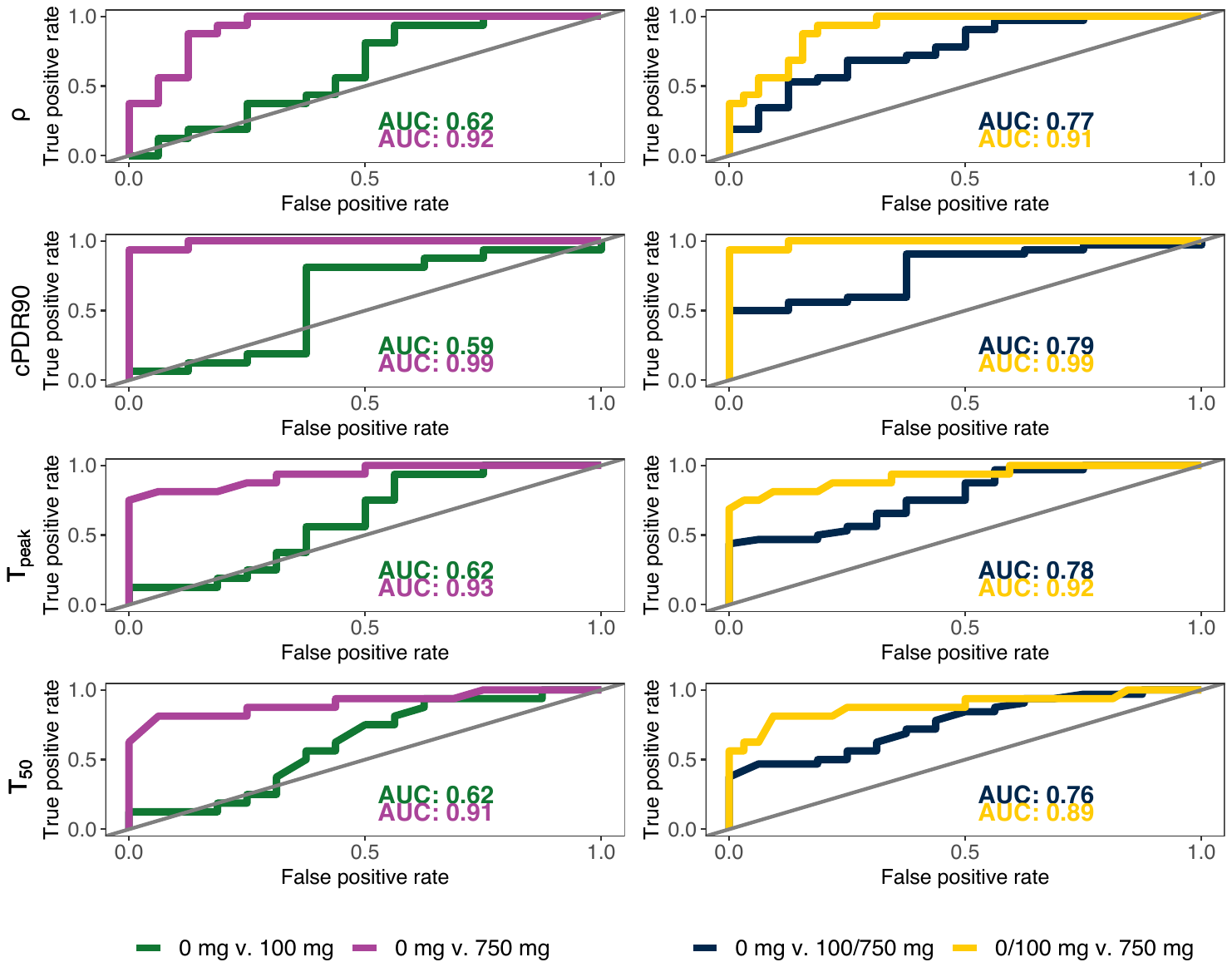

**Figure S4:** ROC curves assuming the data is available at 15 min for hours 0-1, every 30 min for hours 1-4.
